# Neurometabolite Changes Following Football Heading: A 7 T Magnetic Resonance Spectroscopy Study

**DOI:** 10.64898/2026.11.25.26368938

**Authors:** Harshal Jayeshkumar Patel, Syed Ali Muqtadir, Chang-Hoon Choi, Usman Jawed Shaikh, Ana-Maria Oros-Peusquens, Nadim Jon Shah, Colin N Moran, Magdalena Ietswaart, Ferdinand Binkofski

**Affiliations:** Division of Clinical Cognitive Sciences, Medical Faculty of the RWTH Aachen University, Aachen, Germany; School of Psychology, University of Stirling, Stirling, UK; Institute of Neuroscience and Medicine-4, Forschungszentrum Jülich GmbH, Jülich, Germany; Centre for Research and Innovation in Sport, University of Stirling, Stirling, UK; JARA-BRAIN-Translational Medicine, Jülich-Aachen-Research-Alliance (JARA), Aachen, Germany

**Keywords:** traumatic brain injury, repeated subconcussive head impact, concussion, sport, magnetic resonance spectroscopy, soccer

## Abstract

**Importance:** There is growing concern around the acute and cumulative effects of repeated non-concussive head impacts with football heading. However, it is still unclear whether a single bout of heading alters neurochemistry.

**Objective:** To determine whether an acute bout of controlled football heading alters neurometabolite levels measured with 7 T proton magnetic resonance spectroscopy with the primary motor cortex.

**Design:** A within-subject, repeated measures study with proton spectra acquired at three timepoints: one before heading (Pre) and twice after heading (Post_1 and Post_2).

**Setting:** Data were collected using the 7 T human MRI system. Controlled football heading exposure was carried out using an automated football launcher.

**Participants:** 20 participants (15 male), with a mean age of 25.2 ± 6.6 were recruited. Participants had no reported history of head injury in the past year or any neurological conditions.

**Exposure:** A commercial ball launcher was used to deliver 20 controlled headers to each participant standing 10 metres away. The participant remained standing as they executed rotational headers, redirecting the ball towards the right side of the field.

**Main Outcome(s) and Measure(s):** Neurometabolite absolute molal concentrations (mol/Kg).

**Results:** *N-*acetylaspartate (NAA) showed a significant effect of Time [*F*(2, 36.7) = 8.01, *p*_FDR_ < 0.01] with elevated levels at the Post_2 timepoints compared with both Pre and Post_1 measurements. No other metabolites showed significant changes.

**Conclusions and Relevance:** An acute bout of 20 headers was associated with detectable changes in NAA concentration levels. This finding from 7 T MRS provides valuable insight into the neurochemical changes resultant from repeated non-concussive head impacts, which may be suggestive of an acute neuronal response to heading.

**Question:** How does football heading acutely affect neurometabolites over time as measured with ultra-high field magnetic resonance spectroscopy?

**Findings:** In this exploratory study with 20 healthy adults, we detected an increase in *N-*acetylaspartate concentrations within the primary motor cortex at around 90 minutes following heading with no detected acute changes in GABA, glutamate or glutamine.

**Meaning:** Our study points to acute neurometabolite alterations associated with heading, with observed time-dependent NAA elevations.

## Introduction

The recent restrictions on football heading put in place by a mainstream Italian Serie A football club, Como 1907, has put the spotlight back on the safety of football heading.^1^ Combined with the recent deaths of professional footballers attributed to heading-related sequelae,^2,3^ there is significant public interest in the question of heading safety. Footballers have been shown to be at a significantly greater risk for developing neurodegenerative conditions compared to the normal population.^4,5^ This includes chronic traumatic encephalopathy (CTE) even with a minimal or no history of concussion.^6,7^ Despite a preponderance of epidemiological evidence, the acute effects of heading remain less clear with heterogeneity in levels of heading exposure, measured outcomes, and experimental design contributing to this lack of clarity.^8–10^

The excitation-inhibition (EI) balance is one possible framework by which heading may contribute to the chronic pathological processes detailed in the epidemiological studies remarked on previously. A balance between glutamate (Glu)- and gamma-aminobutyric acid (GABA)-mediated excitation and inhibition is necessary for optimal functioning of cellular mechanisms regulating neuronal activity.^11^ Disturbance of this EI balance has been observed following brain injury with alterations in Glu and GABA mediated signalling that can develop from short-term effects into long-term consequences such as deficit in cognitive and motor functions.^12^ There is evidence of non-concussive head impacts can affect EI balance in the form of an acute elongation of the cortical silent period (cSP) - a transcranial magnetic stimulation (TMS) measure that is associated with GABA-B receptor activity within M1.^13^ Changes in GABAergic transmission have also been shown to be time-dependent, showing up one hour after a sparring session in study with boxers.^14^ While these findings are suggestive of transient, time-dependent functional changes being imparted by non-concussive head impacts, it does not speak to any changes that may take place within the neurochemical makeup given that TMS and MRS index different aspects of the EI balance.^15,16^ To the best of our knowledge, no attempt has been made to quantify EI neurometabolite concentration changes before and after a non-concussive heading intervention.

Repeated non-concussive head impacts have also been associated with cellular changes including disruption of the blood-brain barrier,^17^ astrocytic damage^18^ and neuronal degeneration.^19^ Few studies have used *in vivo* proton magnetic resonance spectroscopy (MRS) to investigate heading effects, with fewer still looking at acute effects and none at 7 T. Elevated total *N-*acetylaspartate (tNAA) and total creatine (tCr) levels have been observed 40 minutes after exposure to 20 linear headers in the primary motor cortex (M1), attributed to increased mitochondrial metabolism.^20^ A longitudinal study following female football players found increased M1 glutamate + glutamine (Glx) in the second half of a competitive season and at a 4-8 week post-season follow-up.^21^ In a recent study with adolescent male football players, higher tNAA baseline concentrations in periventricular white matter were observed relative to non-contact athletes which normalised over the course of the season.^22^ Data from a cross-sectional study have shown lower frontal NAA/Cr and higher Glx/Cr levels in a cohort of active football players compared to healthy controls.^23^ Additionally, retired professional football players have been found to have higher choline (Cho) and myo-Inositol (mI) levels compared to non-contact athletes.^24^ Only one of the aforementioned studies has used an interventional design, making it difficult to establish a causative relationship between heading and neurometabolite alternations.

^1^H MRS at 7 T is able to quantify discrete metabolites more precisely and with better quantification compared to lower field strengths.^25^ Ultra-high field spectroscopy also has the advantage of enabling better separation of overlapping peaks like that of NAA and *N*-acetylaspartylglutamate (NAAG).^26^ In this study, we aimed to determine whether exposure to a series of controlled heading events resulted in changes to GABA-mediated inhibition and glutamate-mediated excitation within the M1 over two acute timepoints. Our secondary goal was to observe whether heading-related perturbations were observed in metabolites associated with mechanisms outside EI balance, namely, NAA-related metabolites, energy metabolism and membrane turnover. This exploratory study is the first to employ ultra-high field 7 T MRS in an experimental approach to observing heading effects at two different time points.

## Methods

### Participants and procedure

20 healthy participants (15 male), aged 25.2 ± 6.6 years with no known history of head injury in the past 12 months or neurological conditions were enrolled in the study. The sample size was dictated by time and resource constraints. Written consent was obtained from all participants with approval from the ethics committee at the Faculty of Medicine, RWTH Aachen University, Aachen, Germany. All data collection procedures aligned with the Declaration of Helsinki.

Following an MR safety screening, participants were scanned at three timepoints: before heading (Pre), 40.26 ± 8.19 minutes after heading (Post_1), and 89.67 ± 11.24 minutes after heading (Post_2).

The heading protocol comprised exposure to 20 football headers with a regulation size football inflated to eight psi. Participants were asked to head a ball, projected from a ball launcher (JUGS Soccer Machine; JUGS Sports, Tualatin, OR, USA) positioned 10 m away, on the right side of the field. Participants were led to an open area near the scanner to carry out heading protocol. Participants were instructed to remain standing and avoid excessive movement, including jumping, while attempting the headers. This was done to limit physical activity as a confound. The ball speed at launch was initially set to 26 mph and slightly adjusted according to participant height and comfort. Two experimenters verified each header impact visually, with an impact sensor, and with a participant rating of the head-to-ball contact. The experimental protocol is visualised in Figure 1.

**Figure 1.**
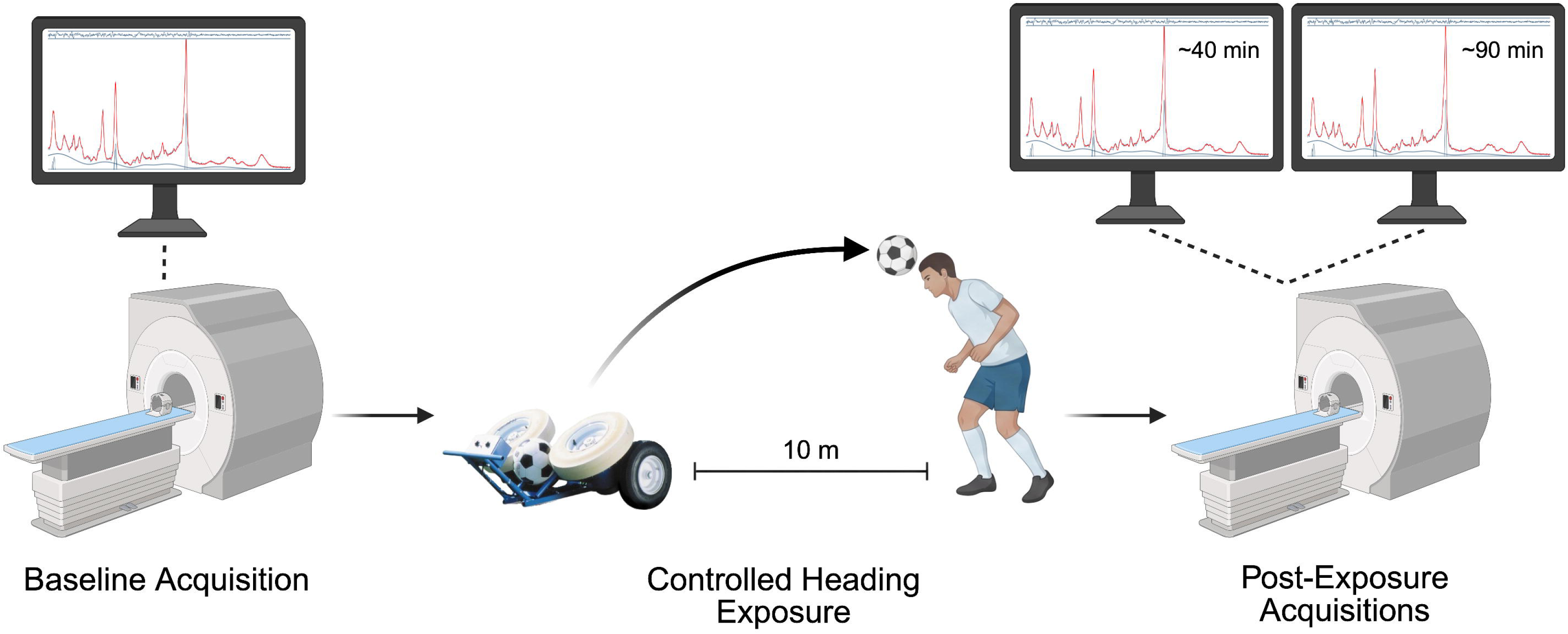
Experimental protocol. MRS spectra were acquired once prior to and twice after (at ∼40 min and at ∼90 min) after controlled heading exposure. Figure created with BioRender.com

Heading impact kinematics were quantified using a custom skin-patch accelerometer attached to the right mastoid and recorded as peak linear acceleration (PLA; *g*) and peak angular acceleration (PRA; rad/s^2^).

### MR acquisition and processing

Participants were scanned with a 7 T Magnetom Terra scanner (Siemens Healthineers, Erlangen, Germany) using a single transmit and 32-channel receive array (Nova Medical, Wilmington, MA, USA). A magnetization-prepared rapid acquisition gradient echo (MP2RAGE) structural image was acquired at the start of both Pre and Post_1 measurements [isotropic resolution = 0.6 × 0.6 × 0.6 mm^3^, field of view (FOV) = 240 × 240 mm^2^, matrix size = 400 × 400, repetition time (TR) = 4440 ms, echo time (TE) = 2.08 ms, inversion time 1 (TI_1_) = 840 ms, inversion time 2 (TI_2_) = 2370 ms, number of averages = 1, GRAPPA factor = 3, total acquisition time = 11:03 minutes]. This 3D image was used to place a 24 × 22 × 17 mm^3^ MRS voxel within the left M1 at the start of Pre, Post_1 and Post_2 measurements as shown in Figure 2. Advanced shimming using FASTESTMAP was applied with slightly larger voxel size of 26 × 24 × 19 mm^3^ to ensure homogenous B_0_ in the selected MRS voxel.^27^ A semi-LASER MRS sequence^28^ (TR = 9500 ms, TE = 36 ms, number of averages = 56, spectral width = 6 kHz, datapoints = 2048 and VAPOR water suppression), was used to acquire proton spectra (details in eTable 1).

**Figure 2.**
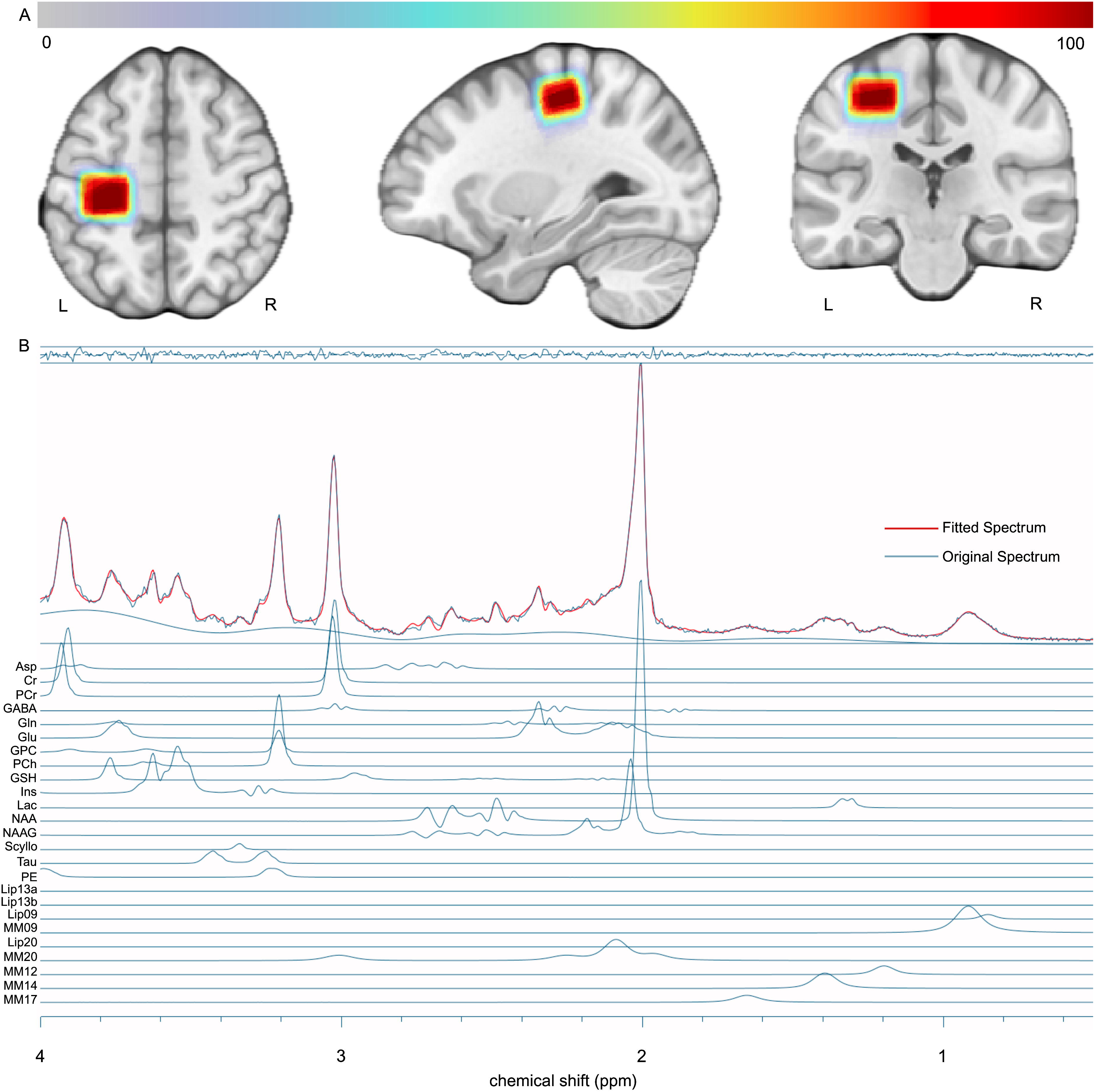
(A) Voxel overlap heatmap generated in Osprey through co-registration of voxels from both Pre and Post acquisitions across participants to a common template space. Warmer colours indicate greater overlap. (B) Sample spectrum from a representative dataset including relative contributions from each metabolite. Fitted spectrum is in red, original MR spectrum is in blue. Fit residual is displayed in the top bar.

MRS analysis was carried out by SAM and independently verified by HJP. A group of individual spectra was exported in coil-combined IMA format for analysis in the open-source Osprey 2.9.6 package running in MATLAB R2023b.^29,30^ Spectral fitting was carried out with LCModel (v6.3-1N). Voxel tissue segmentation was done in Osprey using SPM12. Tissue- and relaxation-corrected molal concentration estimates were calculated in Osprey using the Gasparovic method.^30,31^ Voxel overlap and sample spectrum can be seen in Figure 2 with averaged spectra across timepoints in eFigure 1.

### Statistical analysis

Statistical analyses were carried out in R (version 4.6.0) with lme4, emmeans, and lmerTest packages.^32–35^ Observations with relative Cramér-Rao lower bounds (CRLB) value of 999% were excluded from the analysis. A relative CRLB threshold was not used to exclude observations to avoid biased selection.^36,37^ Rather, the absolute CRLB values were calculated as follows.

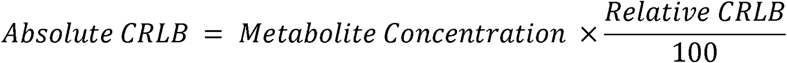

The third quartile (Q3) and interquartile range (IQR) for absolute CRLB at the Pre timepoint were then resolved to create a threshold for each metabolite, defined as *Q*3 + 1.5 *IQR*. Absolute CRLB values from the Post_1 and Post_2 timepoints were then tested against this threshold. Observations were excluded if their values were more than the threshold. For surviving observations, linear mixed models were fitted for each metabolite with a fixed effect of Time (three levels: Pre, Post_1 and Post_2) and a participant random intercept. Where omnibus tests were significant, post-hoc pairwise differences were estimated using estimated marginal (EM) means. For the readers’ interest, the data were also analysed with the commonly used 20% relative CRLB threshold.^38–40^ This did not change the inferences drawn from the results (e2).

Measured metabolites were assigned to four families defined by a mechanism to structure the analysis.^41–43^ These families included the primary EI balance family (GABA, Glu, Gln) with secondary families comprising NAA-related metabolites [NAA, NAAG], energy metabolism [creatine (Cr), phosphocreatine (PCr), lactate (Lac), aspartate (Asp)], membrane turnover (glycerophosphocholine (GPC), phosphocholine (PCh), phosphoethanolamine (PE)]. Within each metabolite family, omnibus p-values were adjusted for multiple comparisons within each family using false discovery rate (FDR). Alpha was set at an FDR-adjusted *p*_FDR_ < 0.05 (two-tailed). Remainder metabolites (myo-Inositol, scyllo-inositol, glutathione, taurine) were reported uncorrected. To see whether any metabolite changes scaled with heading exposure, the cumulative peak linear (cPLA) and rotational (cPRA) accelerations were calculated and converted into z-scores. They were then added to separate linear mixed models as a Time × cPLA/cPRA interaction.

The analysed sample included all scanned 20 individuals. Demographic data are presented in Table 1. Kinematic data were absent for four participants due to technical faults. Following quality control checks, one value for Scyllo was excluded for relative CRLB = 999%. NAAG and Lac had the most observations excluded with 8.3% exceeding the absolute CRLB threshold out of 60 total observations across participants and timepoints. Details for excluded values across metabolites can be found in eTable 3.

**Table 1.** Demographic and session data.

| **Variable** | **Mean ± SD** |
|---|---|
| Age (years) | 25.2 ± 6.6 |
| Peak linear acceleration (g) | 13.18 ± 2.5 |
| Peak rotational acceleration (rad/s<sup>2</sup>) | 674.3 ± 213.2 |
| Participant header impact rating (maximum 10) | 8.03 ± 0.7 |

## Results

Friedman’s tests showed no differences in spectral quality measures between all timepoints (all *p* > 0.82). Wilcoxon tests showed no change in tissue fractions between from Pre to Post (all *p* > 0.87; Table 2). Peak linear and rotational accelerations recorded were 13.18 ± 2.5 *g* and 674.3 ± 213.2 rad/s^2^ respectively. Participants reported good contact with the ball during heading actions with mean contact ratings of 8.03 ± 0.7 out of 10.

**Table 2.**
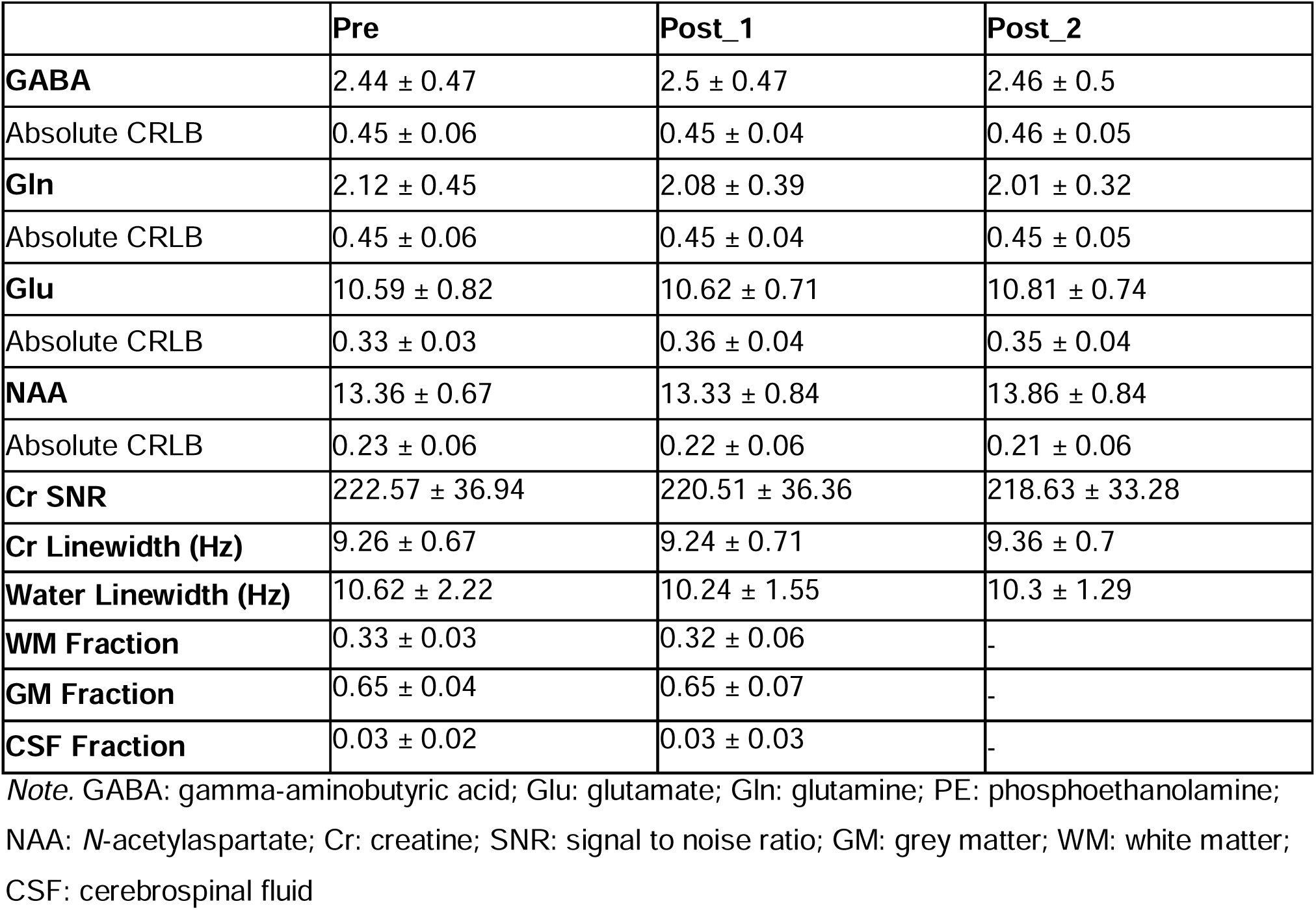
Metabolite absolute molal concentrations (mol/Kg) with associated absolute CRLB values, quality control measures and tissue fractions across timepoints.

|  | **Pre** | **Post_1** | **Post_2** |
|---|---|---|---|
| **GABA** | 2.44 ± 0.47 | 2.5 ± 0.47 | 2.46 ± 0.5 |
| Absolute CRLB | 0.45 ± 0.06 | 0.45 ± 0.04 | 0.46 ± 0.05 |
| **Gln** | 2.12 ± 0.45 | 2.08 ± 0.39 | 2.01 ± 0.32 |
| Absolute CRLB | 0.45 ± 0.06 | 0.45 ± 0.04 | 0.45 ± 0.05 |
| **Glu** | 10.59 ± 0.82 | 10.62 ± 0.71 | 10.81 ± 0.74 |
| Absolute CRLB | 0.33 ± 0.03 | 0.36 ± 0.04 | 0.35 ± 0.04 |
| **NAA** | 13.36 ± 0.67 | 13.33 ± 0.84 | 13.86 ± 0.84 |
| Absolute CRLB | 0.23 ± 0.06 | 0.22 ± 0.06 | 0.21 ± 0.06 |
| **Cr SNR** | 222.57 ± 36.94 | 220.51 ± 36.36 | 218.63 ± 33.28 |
| **Cr Linewidth (Hz)** | 9.26 ± 0.67 | 9.24 ± 0.71 | 9.36 ± 0.7 |
| **Water Linewidth (Hz)** | 10.62 ± 2.22 | 10.24 ± 1.55 | 10.3 ± 1.29 |
| **WM Fraction** | 0.33 ± 0.03 | 0.32 ± 0.06 | - |
| **GM Fraction** | 0.65 ± 0.04 | 0.65 ± 0.07 | - |
| **CSF Fraction** | 0.03 ± 0.02 | 0.03 ± 0.03 | - |

*Note.* GABA: gamma-aminobutyric acid; Glu: glutamate; Gln: glutamine; PE: phosphoethanolamine; NAA: *N*-acetylaspartate; Cr: creatine; SNR: signal to noise ratio; GM: grey matter; WM: white matter; CSF: cerebrospinal fluid

### No changes in GABA, Glu or Gln

No significant change was found for GABA, *F*(2, 36.9) = 0.34, *p*_FDR_ = 0.71, glutamate, *F*(2, 33.96) = 0.99, *p*_FDR_ = 0.65, or Gln, *F*(2, 37.07) = 0.86, *p*_FDR_ = 0.65, metabolites (Table 2; Figure 3).

**Figure 3.**
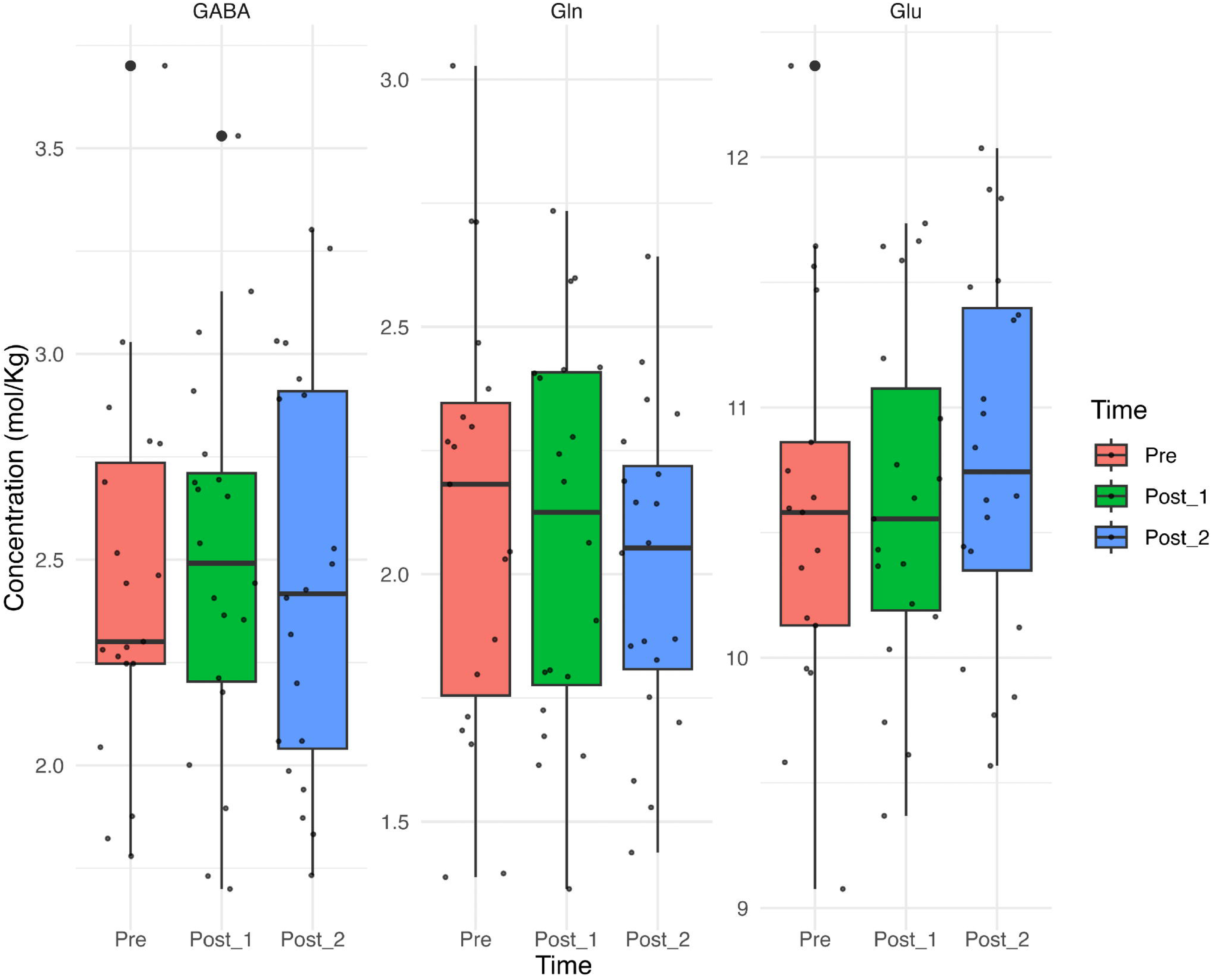
Concentrations (mol/Kg) for GABA, glutamine (Gln), and glutamate (Glu) across all timepoints.

### Increase in NAA ∼90 minutes after heading

An increase was observed in NAA [Time: *F*(2, 36.7) = 8.01, *p*_FDR_ < 0.01; Table 2; Figure 4] with EM means of 13.4 ± 0.18 (Pre), 13.3 ± 0.18 (Post_1), and 13.9 ± 0.18 (Post_2). Post hoc comparisons showed significantly higher concentrations at Post_2 compared to Pre (Δ = 0.43 mol/Kg, 95% CI [0.09, 0.79]*, t*(37.2) = 3.07, *p* < 0.05) and also significantly higher concentrations from Post_1 to Post_2 (Δ = 0.52 mol/Kg, 95% CI [0.18, 0.87]*, t*(37) = 3.75, *p* < 0.01). The change from Pre to Post_1 was not significant (Δ = -0.09 mol/Kg, *p* = 0.81). No significant changes were observed in any of the other metabolites (Figure 5; Table 3). Adding kinematic data from the skin-patch sensor to the model did not show any significant moderating effects of cPLA (*p* = 0.20) or cPRA (*p* = 071). Individual trajectories of concentrations changes from pre to post timepoints for each metabolite is visualised in eFigure 2.

**Figure 4.**
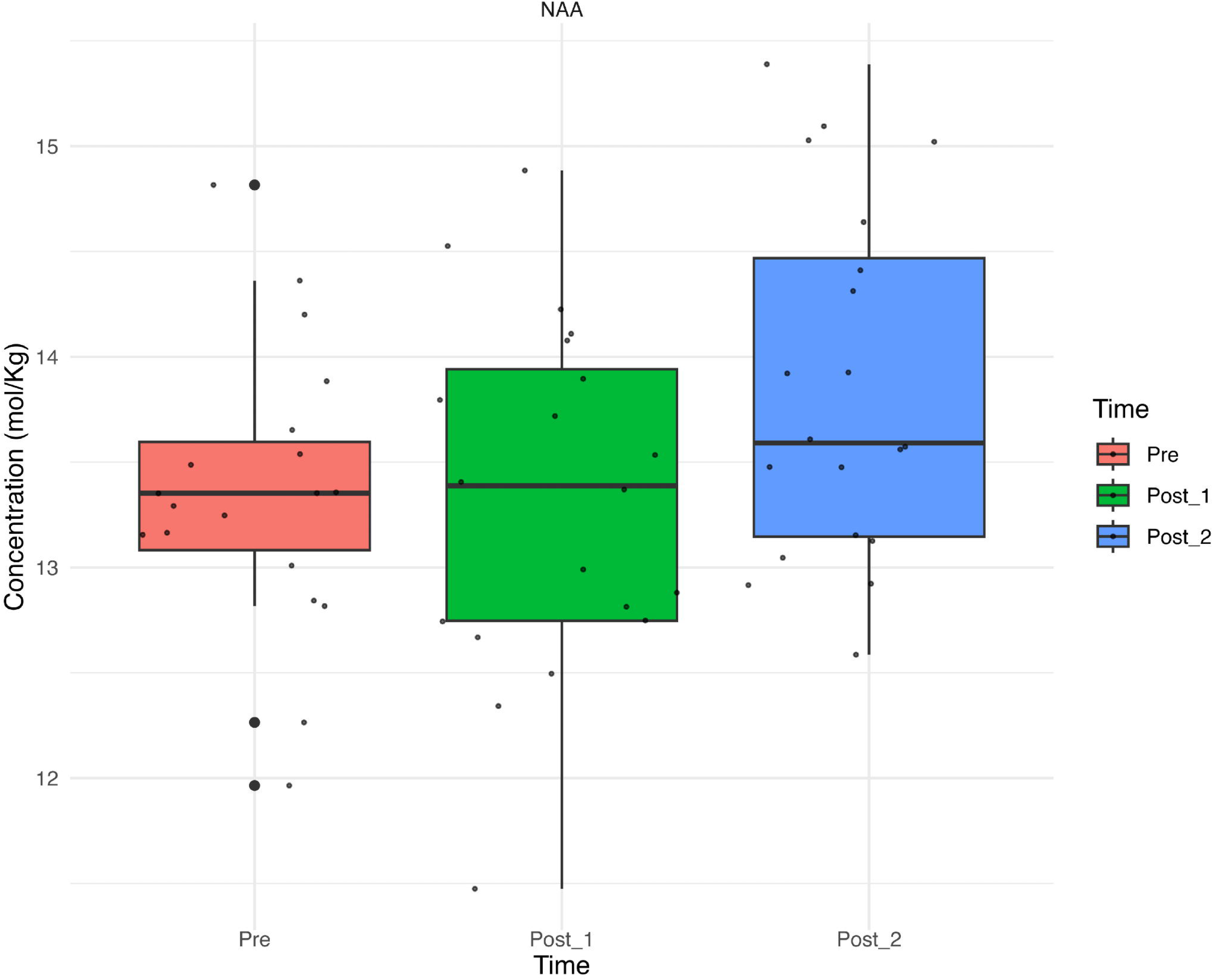
Concentrations (mol/Kg) for *N*-acetylaspartate (NAA) across all timepoints.

**Figure 5.**
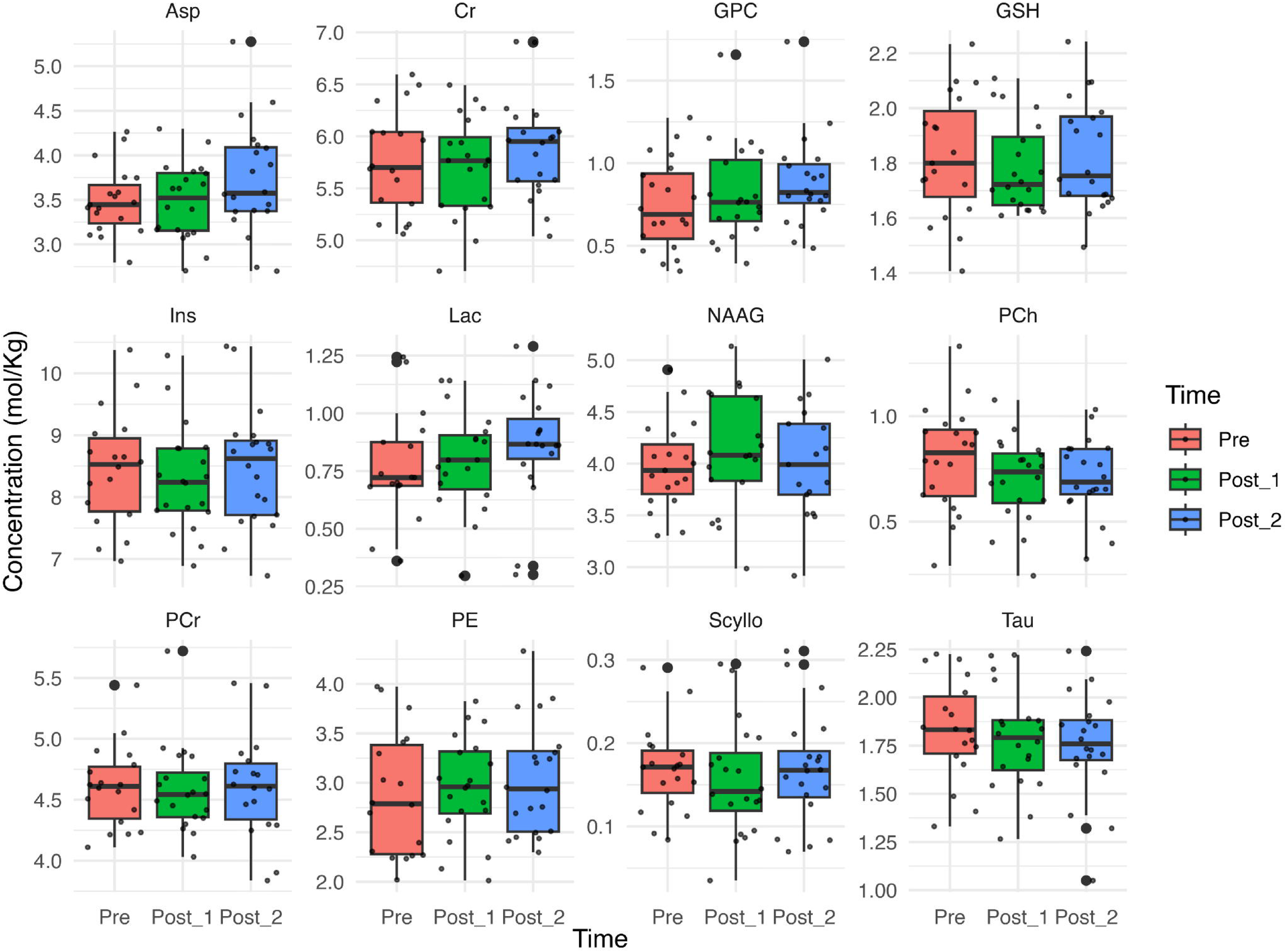
Remainder metabolite concentrations (mol/Kg) across all timepoints.

**Table 3.** Remainder metabolite concentrations (mol/Kg) and absolute CRLB values (Mean ± SD)

|  | **Pre** | **Post_1** | **Post_2** |
|---|---|---|---|
| **Aspartate (Asp)** | 3.49 ± 0.38 | 3.48 ± 0.43 | 3.72 ± 0.63 |
| Absolute CRLB | 0.58 ± 0.06 | 0.58 ± 0.04 | 0.58 ± 0.05 |
| **Creatine (Cr)** | 5.77 ± 0.5 | 5.71 ± 0.48 | 5.88 ± 0.49 |
| Absolute CRLB | 0.19 ± 0.02 | 0.21 ± 0.02 | 0.2 ± 0.03 |
| **Glycerophosphocholine (GPC)** | 0.75 ± 0.27 | 0.82 ± 0.29 | 0.89 ± 0.28 |
| Absolute CRLB | 0.14 ± 0.03 | 0.15 ± 0.03 | 0.14 ± 0.03 |
| **Glutathione (GSH)** | 1.82 ± 0.22 | 1.78 ± 0.16 | 1.83 ± 0.2 |
| Absolute CRLB | 0.12 ± 0.01 | 0.12 ± 0.01 | 0.12 ± 0.01 |
| **Myo-inositol (Ins)** | 8.44 ± 0.92 | 8.27 ± 0.84 | 8.48 ± 0.96 |
| Absolute CRLB | 0.26 ± 0.03 | 0.26 ± 0.03 | 0.26 ± 0.03 |
| **Lactate (Lac)** | 0.77 ± 0.24 | 0.8 ± 0.21 | 0.86 ± 0.24 |
| Absolute CRLB | 0.16 ± 0.01 | 0.17 ± 0.01 | 0.17 ± 0.02 |
| **N-acetylaspartylglutamate (NAAG)** | 3.98 ± 0.42 | 4.12 ± 0.56 | 4.01 ± 0.53 |
| Absolute CRLB | 0.15 ± 0.03 | 0.15 ± 0.03 | 0.14 ± 0.03 |
| **Phosphocholine (PCh)** | 0.8 ± 0.24 | 0.7 ± 0.2 | 0.71 ± 0.18 |
| Absolute CRLB | 0.15 ± 0.03 | 0.16 ± 0.03 | 0.15 ± 0.03 |
| **Phosphocreatine (PCr)** | 4.6 ± 0.34 | 4.59 ± 0.36 | 4.61 ± 0.43 |
| Absolute CRLB | 0.19 ± 0.02 | 0.2 ± 0.02 | 0.2 ± 0.02 |
| **PE** | 2.88 ± 0.63 | 2.95 ± 0.51 | 3.04 ± 0.57 |
| Absolute CRLB | 0.41 ± 0.04 | 0.42 ± 0.03 | 0.43 ± 0.04 |
| **Scyllo-inositol (Scyllo)** | 0.17 ± 0.05 | 0.16 ± 0.07 | 0.17 ± 0.07 |
| Absolute CRLB | 0.04 ± 0.0038 | 0.04 ± 0.0026 | 0.04 ± 0.0029 |
| **Taurine (Tau)** | 1.83 ± 0.26 | 1.78 ± 0.26 | 1.75 ± 0.27 |
| Absolute CRLB | 0.21 ± 0.02 | 0.21 ± 0.01 | 0.21 ± 0.02 |

*Note.* CRLB: Cramer-Rao Lower Bound

## Discussion

Using an experimental approach with a controlled heading intervention, we investigated the effect of repeated non-concussive head impacts on 7 T MRS metabolites over two time points after football heading. We found statistically significant time-dependent changes in a metabolite broadly associated with neuronal health. However, no detectable concentration changes were found in metabolites associated with EI balance (GABA, Gln, and Glu).

While heading has been noted to transiently modulate GABA-B receptor-driven inhibition, changes in the total amount of GABA present within a voxel placed in the M1 were not detected.^13^ This does not preclude the possibility of heading influencing EI balance given the disconnect between ^1^H MRS GABA and GABA-mediated physiological inhibition.^44^ Increased Glx within the M1 has been observed selectively in female football players in the latter half of a competition season and 20-24 week after.^21^ This suggests that while a single bout of heading may not acutely produce measurable EI-related changes, cumulative exposure could have such an effect.

Statistically significant changes in NAA were observed with the effect not appearing to be driven by changes in spectra quality and tissue fractions across measurements. It should be noted that while NAA is generally used a marker for neuronal integrity, it associated with other mechanisms including energy metabolism and lipid synthesis.^41,45^ NAA is synthesised from aspartate and acetyl coenzyme-A (CoA) by an NAA synthase, N-acetyltransferase-8 like protein (NAT8L) with brain injury modulating NAA and acetyl CoA levels in animal models.^46^ NAA is a precursor for NAAG, which is a neuroactive peptide involved in modulation neurotransmitter release.^47^ Increased NAA correlates with increased NAAG levels which, through experimental manipulation, has been found to have neuroprotective effects against excitotoxic effects.^45^ It has also been suggested that, in response to brain injury, NAA may act to support oligodendrocyte metabolism where it is catabolised to form acetate which in turn involved in injury-response mechanisms and myelin sheath repair.^45^ It is uncertain which mechanism drove observed increase in NAA. Furthermore, although NAA showed a significant increase at the group level at the later timepoint, individual responses were variable. This suggests that the acute metabolic response to a uniform heading exposure is not homogenous across participants.

Within mild traumatic brain injury literature, NAA is found to consistently decrease post-injury, with the largest reductions being observed in the hyperacute phase.^48^ It should be noted that this time scale is in the order of days rather than in hours and minutes.^49–51^ Nevertheless, a similar pattern was found in meta-analysis of reports investigating the effect of subconcussive impacts on ^1^H MRS metabolites with reductions in NAA and total creatine (tCr) observed over the course of a season.^52^ While these effects were reported as not stable, the data presented also came from longitudinal studies which are unable to interrogate any causal links between non-concussive impacts and NAA temporal dynamics nor, by design, are able to speak to any changes that may take place over the immediate time course.

The direction of the present NAA result mirrors the acute increase in tNAA (NAA + NAAG) found by Delang et al.^20^ 58 ± 7 minutes after 20 headers within the M1 using 3 T MRS. Of note here is the time differential as well as the fact that the present work, at a higher field strength, did not detect any changes in NAAG. Taken together, the observed tNAA increase appear to be driven primarily by NAA and if this assumption holds true then NAA concentration changes would appear to be detectable between ∼40 and 60 minutes after heading exposure and remain so until at least ∼90 minutes post exposure.

Koerte et al.^22^ recently reported higher baseline levels of tNAA at 3 T in football youth players compared to controls which converged over the season. They speculated that the increased baseline levels could be attributed to the sampling and pre-season training being temporally proximate. This would be a reasonable argument as the heading incidence in training significantly exceeds that in match play by about 600 headers per 1000 player hours within youth football.^53^ While NAA appears to show a time-dependent increase following non-concussive impacts, the present sampling window precludes any comment on whether the pattern of decreasing NAA observed within mTBI studies in the days following injury holds with repeated impacts of lesser magnitude.

More work is needed to understand how NAA levels change over time following non-concussive and mTBI exposure. Studies in ice hockey and American football have reported reduction in NAA measured over a season, whereas no significant NAA changes were observed in female rugby athletes following either concussion or a season.^54–57^ These findings suggest that repeated head impacts may influence neuronal metabolism, but they do no establish a consistent direct of NAA change.

They also suggest NAA response to repetitive head impacts are variable and depend on type, frequency and timing of exposure. Most of the previous studies have examined neurometabolic change over relatively long period, such as competitive season or following concussion.

The inferences drawn from this study are constrained by its design. We were only able to capture the acute time window following heading given the resource-intensive nature of ultra-high field MR scanning and did not include a sham condition. Our findings are also limited to the M1, further work is needed to see if the changes observed here are region-specific or distributed. Skin-patch sensors can capture kinematic data and provide peak acceleration values as a proxy for quantifying head impact exposure. However, other modalities such as instrumented mouthguards are better coupled with the skull and can provide more accurate data that can then be fed into biomechanical models for more information on brain deformation and tissue strain.^58–60^

## Conclusion

Findings from this exploratory study show that exposure to repeated non-concussive head impacts can alter neurometabolite levels in a time-dependent fashion. We found elevated NAA levels at ∼90 minutes but not at ∼40 minutes after heading within the left M1. No other metabolites showed statistically significant changes.

Further work needs to be done to establish the temporal trajectory of NAA following exposure to repeated non-concussive head impacts and whether a dose-response relationship exists between NAA and heading exposure.

## Data Availability

All data produced in the present study are available upon reasonable request to the authors.

## Acknowledgements

Ms. Bechholz and Ms. Köth for their measurement support, Dr. Farrher for the technical discussion and support and CMRR for providing the semi-LASER sequence and FASTESTMAP.^27,61–66^ The MRS package was developed by Edward J. Auerbach and Małgorzata Marjańska and provided by the University of Minnesota under a C2P agreement.

## Notes

### Competing Interest Statement

The authors have declared no competing interest.

### Author Declarations

Written consent was obtained from all participants with approval from the ethics committee at the Faculty of Medicine, RWTH Aachen University, Aachen, Germany

## References

1. Como. Como 1907 Academy introduces new youth rules to support brain health and player development. Como 1907. 24 July 2026. Accessed 24 July 2026. https://comofootball.com/en/como-1907-academy-introduces-new-youth-rules-to-support-brain-health-and-player-development/

2. Smyth M. Heading footballs caused Nobby Stiles’ brain disease, coroner finds. 15 July 2026. Accessed 18 August 2026. https://www.bbc.co.uk/news/articles/c5yzyygmxvgo

3. Austin D, Gornall K. Gordon McQueen death: Inquest finds heading ‘likely’ to have contributed to brain disease. 26 January 2026. Accessed 18 August 2026. https://www.bbc.co.uk/sport/football/articles/c0er74njxgro

4. Mackay DF, Russell ER, Stewart K, MacLean JA, Pell JP, Stewart W. Neurodegenerative Disease Mortality among Former Professional Soccer Players. N Engl J Med. 2019;381(19):1801–1808.

5. Russell ER, Mackay DF, Stewart K, MacLean JA, Pell JP, Stewart W. Association of Field Position and Career Length With Risk of Neurodegenerative Disease in Male Former Professional Soccer Players. JAMA Neurol. 2021;78(9):1057–1063.

6. Rodriguez RD, Suemoto CK, Brucki SMD, et al. Multiple pathologies in an amateur soccer player with severe chronic traumatic encephalopathy: Human neuropathology/other. Alzheimers Dement. 2020;16(S2):e046391.

7. Grinberg LT, Anghinah R, Nascimento CF, et al. Chronic traumatic encephalopathy presenting as Alzheimer’s disease in a retired soccer player. J Alzheimers Dis. 2016;54(1):169–174.

8. Mohamed NSA, Marium R, Osman HH, et al. Impact of repetitive soccer heading on brain health outcomes: a narrative review. Brain Struct Funct. 2025;230(6):108.

9. Lember LM, Ntikas M, Mondello S, et al. The Use of Biofluid Markers to Evaluate the Consequences of Sport-Related Subconcussive Head Impact Exposure: A Scoping Review. Sports Med Open. 2024;10(1):12.

10. Herold F, Pontifex MB, Gronwald T, Heath M. Does a header a day keep the positive effects of physical activity on cognitive performance away? A call to consider physical activity-related head impacts as a moderator variable. Sport Sci Health. 2026;22(1):4.

11. Sears SMS, Hewett SJ. Influence of glutamate and GABA transport on brain excitatory/inhibitory balance. Exp Biol Med (Maywood). 2021;246(9):1069–1083.

12. Guerriero RM, Giza CC, Rotenberg A. Glutamate and GABA Imbalance Following Traumatic Brain Injury. Curr Neurol Neurosci Rep. 2015;15(5):27.

13. Di Virgilio TG, Hunter A, Wilson L, et al. Evidence for Acute Electrophysiological and Cognitive Changes Following Routine Soccer Heading. eBioMedicine. 2016;13:66–71.

14. Di Virgilio TG, Ietswaart M, Wilson L, Donaldson DI, Hunter AM. Understanding the Consequences of Repetitive Subconcussive Head Impacts in Sport: Brain Changes and Dampened Motor Control Are Seen After Boxing Practice. Front Hum Neurosci. 2019;13. doi:10.3389/fnhum.2019.00294

15. Tremblay S, Beaulé V, Proulx S, et al. Relationship between transcranial magnetic stimulation measures of intracortical inhibition and spectroscopy measures of GABA and glutamate+glutamine. J Neurophysiol. 2013;109(5):1343–1349.

16. Stagg CJ, Bachtiar V, Johansen-Berg H. What are we measuring with GABA magnetic resonance spectroscopy? Commun Integr Biol. 2011;4(5):573–575.

17. Marchi N, Bazarian JJ, Puvenna V, et al. Consequences of repeated blood-brain barrier disruption in football players. PLoS One. 2013;8(3):e56805.

18. O’Brien WT, Spitz G, Xie B, et al. Biomarkers of neurobiologic recovery in adults with sport-related concussion. JAMA Netw Open. 2024;7(6):e2415983.

19. Rubin LH, Tierney R, Kawata K, et al. NFL blood levels are moderated by subconcussive impacts in a cohort of college football players. Brain Inj. 2019;33(4):456–462.

20. Delang N, Robertson RV, Tinoco Mendoza FA, et al. The acute effects of non-concussive head impacts on brain microstructure, chemistry and function in male soccer players: A pilot randomised controlled trial. Sports Med Open. 2025;11(1):77.

21. Bari S, Svaldi DO, Jang I, et al. Dependence on subconcussive impacts of brain metabolism in collision sport athletes: an MR spectroscopic study. Brain Imaging Behav. 2019;13(3):735–749.

22. Koerte IK, Wiegand TLT, Bonke EM, et al. Youth soccer participation and brain health Outcomes in adolescent athletes. JAMA Netw Open. 2026;9(6):e2619569.

23. Urbanik A, Guz W, Brożyna M, Ostrogórska M. Changes in the central nervous system in football players: an MRI study. Acta Radiol. 2024;65(8):967–974.

24. Koerte IK, Lin AP, Muehlmann M, et al. Altered neurochemistry in former professional soccer players without a history of concussion. J Neurotrauma. 2015;32(17):1287–1293.

25. Platt T, Ladd ME, Paech D. 7 Tesla and beyond: Advanced methods and clinical applications in magnetic resonance imaging: Advanced methods and clinical applications in magnetic resonance imaging. Invest Radiol. 2021;56(11):705–725.

26. Pradhan S, Bonekamp S, Gillen JS, et al. Comparison of single voxel brain MRS AT 3T and 7T using 32-channel head coils. Magn Reson Imaging. 2015;33(8):1013–1018.

27. Gruetter R, Tkác I. Field mapping without reference scan using asymmetric echo-planar techniques. Magn Reson Med. 2000;43(2):319–323.

28. Scheenen TWJ, Klomp DWJ, Wijnen JP, Heerschap A. Short echo time 1H-MRSI of the human brain at 3T with minimal chemical shift displacement errors using adiabatic refocusing pulses. Magn Reson Med. 2008;59(1):1–6.

29. MathWorks. MATLAB Version 23.2.0285933. 2025.

30. Oeltzschner G, Zöllner HJ, Hui SCN, et al. Osprey: Open-source processing, reconstruction & estimation of magnetic resonance spectroscopy data. J Neurosci Methods. 2020;343(108827):108827.

31. Gasparovic C, Song T, Devier D, et al. Use of tissue water as a concentration reference for proton spectroscopic imaging. Magn Reson Med. 2006;55(6):1219–1226.

32. R Core Team. R: A Language and Environment for Statistical Computing. R Foundation for Statistical Computing; 2025. https://www.R-project.org/

33. Bates D, Mächler M, Bolker B, Walker S. Fitting Linear Mixed-Effects Models Using lme4. Journal of Statistical Software. 2015;67(1):1–48. doi:10.18637/jss.v067.i01

34. Lenth RV, Piaskowski J. emmeans: Estimated Marginal Means, aka Least-Squares Means. Preprint posted online 2025. https://rvlenth.github.io/emmeans/

35. Kuznetsova A, Brockhoff PB, Christensen RHB. lmerTest Package: Tests in Linear Mixed Effects Models. Journal of Statistical Software. 2017;82(13):1–26. doi:10.18637/jss.v082.i13

36. Kreis R. The trouble with quality filtering based on relative Cramér-Rao lower bounds. Magn Reson Med. 2016;75(1):15–18.

37. Li M, Demenescu LR, Colic L, et al. Temporal Dynamics of Antidepressant Ketamine Effects on Glutamine Cycling Follow Regional Fingerprints of AMPA and NMDA Receptor Densities. Neuropsychopharmacology. 2017;42(6):1201–1209.

38. Okada T, Kuribayashi H, Kaiser LG, et al. Repeatability of proton magnetic resonance spectroscopy of the brain at 7 T: effect of scan time on semi-localized by adiabatic selective refocusing and short-echo time stimulated echo acquisition mode scans and their comparison. Quant Imaging Med Surg. 2021;11(1):9–20.

39. van de Bank BL, Emir UE, Boer VO, et al. Multi-center reproducibility of neurochemical profiles in the human brain at 7 T: MC7T-747. NMR Biomed. 2015;28(3):306–316.

40. Cheong I, Marjańska M, Deelchand DK, Eberly LE, Walk D, Öz G. Ultra-high field proton MR spectroscopy in early-stage amyotrophic lateral sclerosis. Neurochem Res. 2017;42(6):1833–1844.

41. Rae CD. A guide to the metabolic pathways and function of metabolites observed in human brain 1H magnetic resonance spectra. Neurochem Res. 2014;39(1):1–36.

42. Rae CD, Rowlands BD, Balcar VJ. Aspartate in the brain: A review. Neurochem Res. 2025;50(3):199.

43. Menshchikov PE, Semenova NA, Manzhurtsev AV, Akhadov TA, Varfolomeev SD. Cerebral quantification of N-acetyl aspartate, aspartate, and glutamate levels in local structures of the human brain using J-editing of 1H magnetic resonance spectra in vivo. Russ Chem Bull. 2018;67(4):655–662.

44. Dyke K, Pépés SE, Chen C, et al. Comparing GABA-dependent physiological measures of inhibition with proton magnetic resonance spectroscopy measurement of GABA using ultra-high-field MRI. Neuroimage. 2017;152:360–370.

45. Moffett JR, Arun P, Ariyannur PS, Namboodiri AMA. N-Acetylaspartate reductions in brain injury: impact on post-injury neuroenergetics, lipid synthesis, and protein acetylation. Front Neuroenergetics. 2013;5:11.

46. Vagnozzi R, Tavazzi B, Signoretti S, et al. Temporal window of metabolic brain vulnerability to concussions: mitochondrial-related impairment--part I: Mitochondrial-related impairment-part i. Neurosurgery. 2007;61(2):379–388; discussion 388-9.

47. Benarroch EE. N-acetylaspartate and N-acetylaspartylglutamate: neurobiology and clinical significance. Neurology. 2008;70(16):1353–1357.

48. Joyce JM, La PL, Walker R, Harris AD. Magnetic Resonance Spectroscopy of Traumatic Brain Injury and Subconcussive Hits: A Systematic Review and Meta–Analysis. J Neurotrauma. 2022;39(21-22):1455–1476.

49. Vagnozzi R, Signoretti S, Cristofori L, et al. Assessment of metabolic brain damage and recovery following mild traumatic brain injury: a multicentre, proton magnetic resonance spectroscopic study in concussed patients. Brain. 2010;133(11):3232–3242.

50. Vagnozzi R, Signoretti S, Tavazzi B, et al. Temporal window of metabolic brain vulnerability to concussion: a pilot 1H-magnetic resonance spectroscopic study in concussed athletes--part III: A pilot 1h-magnetic resonance spectroscopic study in concussed athletes-part iii. Neurosurgery. 2008;62(6):1286–1295; discussion 1295–6.

51. Giza CC, Hovda DA. The new neurometabolic cascade of concussion. Neurosurgery. 2014;75 Suppl 4(Supplement 4):S24–33.

52. Delang N, Irwin C, Peek AL, McGregor IS, Desbrow B, McCartney D. The effect of contact/collision sport participation without concussion on neurometabolites: A systematic review and meta-analysis of magnetic resonance spectroscopy studies. J Neurochem. 2023;167(5):615–632.

53. Reeschke R, Haase FK, Dautzenberg L, Krutsch W, Reinsberger C. Training matters: Heading incidence and characteristics in children’s and youth football (soccer) players. Scand J Med Sci Sports. 2023;33(9):1821–1830.

54. Chamard E, Théoret H, Skopelja EN, Forwell LA, Johnson AM, Echlin PS. A prospective study of physician-observed concussion during a varsity university hockey season: metabolic changes in ice hockey players. Part 4 of 4. Neurosurg Focus. 2012;33(6):E4: 1–7.

55. Poole VN, Abbas K, Shenk TE, et al. MR spectroscopic evidence of brain injury in the non-diagnosed collision sport athlete. Dev Neuropsychol. 2014;39(6):459–473.

56. Schranz AL, Manning KY, Dekaban GA, et al. Reduced brain glutamine in female varsity rugby athletes after concussion and in non-concussed athletes after a season of play. Hum Brain Mapp. 2018;39(4):1489–1499.

57. Vike N, Bari S, Susnjar A, et al. American football position-specific neurometabolic changes in high school athletes: A magnetic resonance spectroscopic study. J Neurotrauma. 2022;39(17-18):1168–1182.

58. Huber CM, Patton DA, Maheshwari J, Zhou Z, Kleiven S, Arbogast KB. Finite element brain deformation in adolescent soccer heading. Comput Methods Biomech Biomed Engin. 2024;27(10):1239–1249.

59. Perkins RA, Bakhtiarydavijani A, Ivanoff AE, Jones M, Hammi Y, Prabhu RK. Assessment of brain injury biomechanics in soccer heading using finite element analysis. Brain Multiphys. 2022;3:100052.

60. Liu Y, Domel AG, Yousefsani SA, et al. Validation and comparison of instrumented mouthguards for measuring head kinematics and assessing brain deformation in football impacts. Ann Biomed Eng. 2020;48(11):2580–2598.

61. Klomp DWJ, Bitz AK, Heerschap A, Scheenen TWJ. Proton spectroscopic imaging of the human prostate at 7 T. NMR Biomed. 2009;22(5):495–501.

62. Tremblay S, Beaulé V, Proulx S, et al. The use of magnetic resonance spectroscopy as a tool for the measurement of bi-hemispheric transcranial electric stimulation effects on primary motor cortex metabolism. J Vis Exp. 2014;(93):e51631.

63. Marjańska M, Lehéricy S, Valabrègue R, et al. Brain dynamic neurochemical changes in dystonic patients: a magnetic resonance spectroscopy study: MRS Study in Dystonic Patients After TMS. Mov Disord. 2013;28(2):201–209.

64. Garwood M, DelaBarre L. The return of the frequency sweep: designing adiabatic pulses for contemporary NMR. J Magn Reson. 2001;153(2):155–177.

65. Tkác I, Starcuk Z, Choi IY, Gruetter R. In vivo 1H NMR spectroscopy of rat brain at 1 ms echo time. Magn Reson Med. 1999;41(4):649–656.

66. Allaïli N, Valabrègue R, Auerbach EJ, et al. Single-voxel (1)H spectroscopy in the human hippocampus at 3 T using the LASER sequence: characterization of neurochemical profile and reproducibility: MRS in the Human Hippocampus at 3 T. NMR Biomed. 2015;28(10):1209–1217.

