## Supplementary Material for "Neurometabolite Changes Following Football Heading: A 7 T Magnetic Resonance Spectroscopy Study"

**eTable 1.** Minimum Reporting Standards for *in vivo* Magnetic Resonance Spectroscopy (MRSinMRS)

| **1.** **Hardware** | |
| --- | --- |
| a. Field strength [Tesla] | 7T |
| b. Manufacturer | Siemens Healthineers, Erlangen, Germany |
| c. Model | Magnetom Terra syngo MR E12 |
| d. RF coil | Single-transmit and 32-channel receive coil array |
| e. Additional hardware | N/a |
| **2.** **Acquisition** | |
| a. Pulse sequence | semi-LASER |
| b. Volume of interest (VOI) | single voxel placed in left primary motor cortex |
| c. Nominal VOI size | 24 x 22 x 17 mm^3^ |
| d. Repetition time (TR), echo time (TE) | TR: 9500 ms, TE: 36 ms |
| e. Total number of excitations or acquisitions per spectrum | 56 |
| f. Additional sequence parameters | Spectral bandwidth: 6 kHz, number of points: 2048 |
| g. Water suppression method | VAPOR |
| h. Shimming method | FASTESTMAP |
| i. Triggering or motion correction method | N/a |
| **3.** **Data analysis methods and outputs** | |
| a. Analysis software | Osprey (version 2.9.6), LCModel (version 6.3-1N linked to Osprey), SPM12 |
| b. Processing steps deviating from quoted reference or product | N/A |
| c. Output measure | Absolute molal concentration, using tissue-corrected water (mol/kg; TissCorrWaterScaled as reported by Osprey Quantify function) |
| d. Quantification references and assumptions, fitting model assumptions | Fit range: 0.5-4.0 ppm    Osprey bLineKnotSpace: 0.4  Simulated basis set: aspartate (Asp), creatine (Cr), phosphocreatine (PCr), gamma-aminobutyric acid (GABA), glutamine (Gln), glutamate (Glu), glycerophosphocholine (GPC), phosphocholine (PCh), glutathione (GSH), lactate (Lac), myo-inositol (Ins), *N*-acetylaspartate (NAA), *N*-acetylaspartylglutamate (NAAG), phosphoethanolamine (PE), scyllo-inositol (Scyllo), taurine (Tau) and macromolecules |
| **4.** **Data quality** | |
| a. Reported variables | creatine SNR, creatine linewidth, and water linewidth as reported by Osprey |
| b. Data exclusion criteria | Poor fit or water suppression, absolute CRLB median ± 3 x median absolute deviation |
| c. Quality measures of postprocessing model fitting | Cramer-Rao lower bounds (CRLB) as reported by LCModel |
| d. Sample spectrum | Figure 1 |

*Note.* Reporting table from the consensus paper by Lin et al. (2021).^1^

**eTable 2**. Omnibus results with relative CRLB > 20% filtered data

|  | **Metabolite** | **Statistic** |
| --- | --- | --- |
| **Excitation-inhibition** | Glu | F(2,38) = 1.32, *p*_FDR_ = 0.42 |
|  | GABA | F(2,26.34) = 0.71, *p*_FDR_ = 0.501 |
| **Energy Metabolism** | Asp | F(2,35.5) = 3.26, *p*_FDR_ = 0.201 |
|  | Cr | F(2,38) = 1.94, *p*_FDR_ = 0.314 |
|  | PCr | F(2,38) = 1.1, *p*_FDR_ = 0.456 |
| **Membrane Turnover** | PCh | F(2,20.12) = 3.31, *p*_FDR_ = 0.172 |
|  | GPC | F(2,25.16) = 1.24, *p*_FDR_ = 0.46 |
|  | PE | F(2,34.44) = 0.64, *p*_FDR_ = 0.533 |
| **NAA-related metabolites** | NAA | F(2,38) = 5.98, *p*_FDR_ = 0.011 |
|  | NAAG | F(2,38) = 1.46, *p*_FDR_ = 0.244 |
| **Others** | Ins | F(2,38) = 1.99, *p* = 015 |
|  | Tau | F(2,38) = 1.73, *p* = 0.191 |
|  | GSH | F(2,38) = 0.82, *p* = 0.45 |

*Note.* Scyllo, Gln, and Lac with mean ± SD relative CRLBs of 27.8 ± 15.4, 23 ± 5.9, and 24 ± 9.7 respectively, had more than 50% of observations excluded.

**eTable 3**. Quality control output

| **Metabolite** | **Relative CRLB** | **Absolute CRLB** | **Percentage Excluded** |
| --- | --- | --- | --- |
| Lac | 0 | 5 | 8.3 |
| NAAG | 0 | 5 | 8.3 |
| Glu | 0 | 4 | 6.7 |
| PCr | 0 | 4 | 6.7 |
| Cr | 0 | 2 | 3.3 |
| Ins | 0 | 2 | 3.3 |
| PE | 0 | 2 | 3.3 |
| Tau | 0 | 2 | 3.3 |
| Asp | 0 | 1 | 1.7 |
| GABA | 0 | 1 | 1.7 |
| GSH | 0 | 1 | 1.7 |
| Gln | 0 | 1 | 1.7 |
| NAA | 0 | 1 | 1.7 |
| GPC | 0 | 0 | 0 |
| PCh | 0 | 0 | 0 |
| Scyllo | 1 | 0 | 0 |

*Note.* Total number of observations = 60, Relative CRLB: Values excluded due to being equal to 999%, Absolute CRLB: Values excluded due to being more than the predefined threshold.

**eFigure 1**

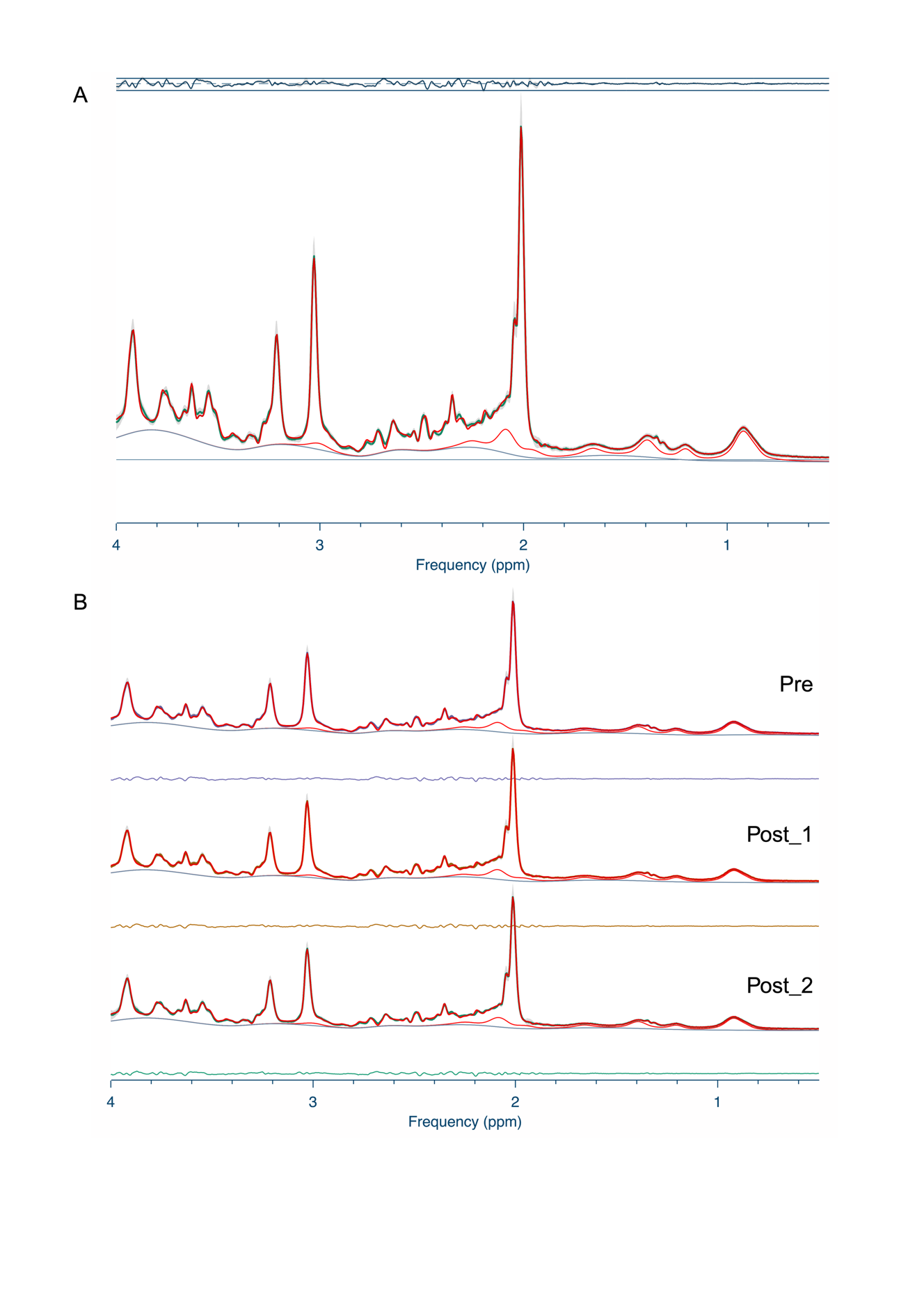

**eFigure 2**

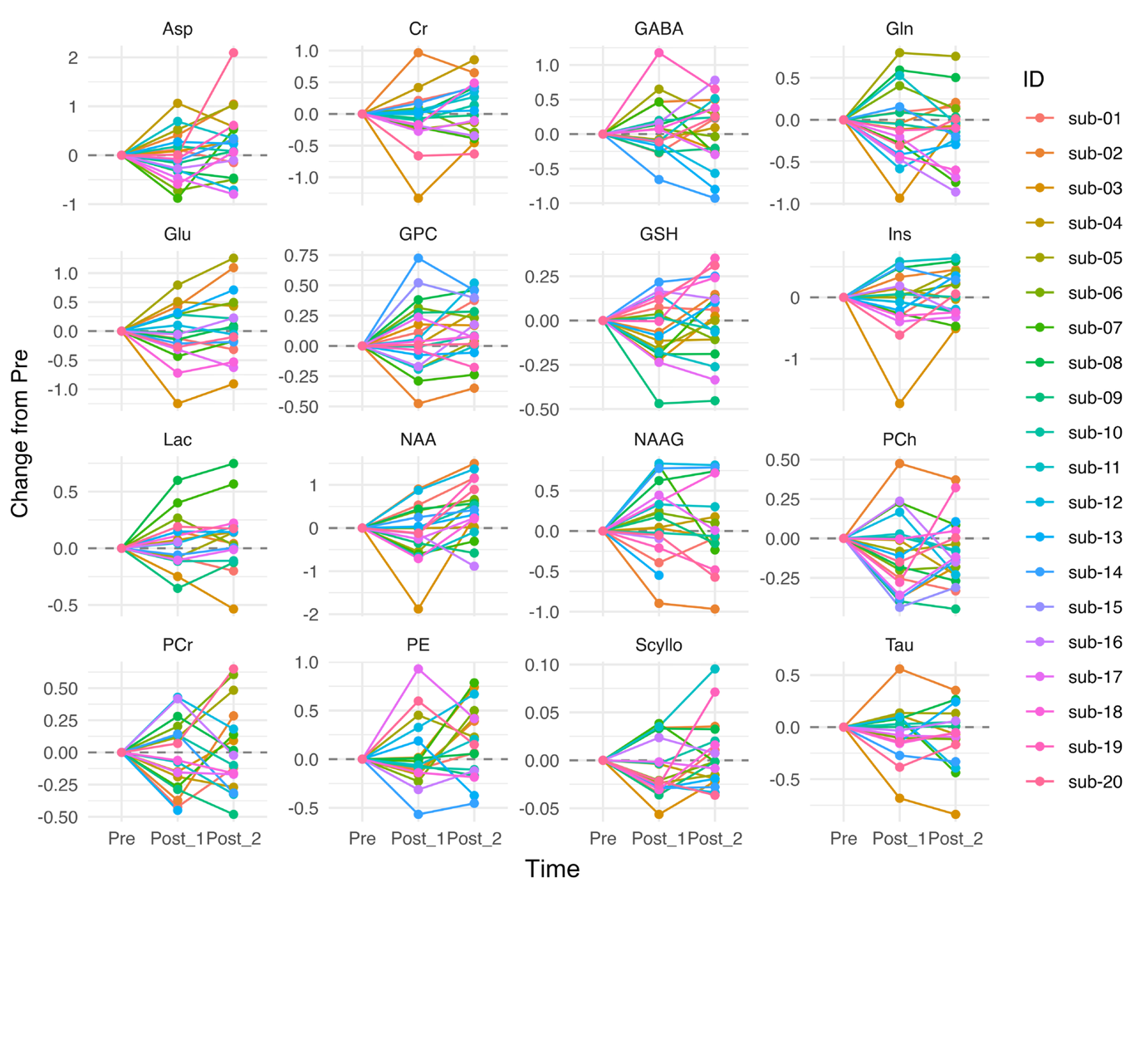

**References**

1.	Lin A, Andronesi O, Bogner W, et al. Minimum Reporting Standards for in vivo Magnetic Resonance Spectroscopy (MRSinMRS): Experts’ consensus recommendations. *NMR Biomed*. 2021;34(5):e4484.
